# Benzodiazepine Initiation and the Risk of Falls or Fall-Related Injuries in Older Adults Following Acute Ischemic Stroke

**DOI:** 10.1101/2024.02.06.24302430

**Authors:** Shuo Sun, Victor Lomachinsky, Louisa H. Smith, Joseph P. Newhouse, M. Brandon Westover, Deborah Blacker, Lee Schwamm, Sebastien Haneuse, Lidia M.V.R. Moura

## Abstract

**Background:** Benzodiazepine use in older adults following acute ischemic stroke (AIS) is common, yet short-term safety concerning falls or fall-related injuries remains unexplored.

**Methods:** We emulated a hypothetical randomized trial of benzodiazepine use during the acute post stroke recovery period to assess incidence of falls or fall related injuries in older adults. Using linked data from the Get With the Guidelines Registry and Mass General Brigham’s electronic health records, we selected patients aged 65 and older admitted for Acute Ischemic Stroke (AIS) between 2014 and 2021 with no documented prior stroke and no benzodiazepine prescriptions in the previous 3 months. Potential for immortal-time and confounding biases was addressed via separate inverse-probability weighting strategies.

**Results:** The study included 495 patients who initiated inpatient benzodiazepines within three days of admission and 2,564 who did not. After standardization, the estimated 10-day risk of falls or fall-related injuries was 694 events per 1000 (95% confidence interval CI: 676-709) for the benzodiazepine initiation strategy and 584 events per 1000 (95% CI: 575-595) for the non-initiation strategy. Subgroup analyses showed risk differences of 142 events per 1000 (95% CI: 111-165) and 85 events per 1000 (95% CI: 64-107) for patients aged 65 to 74 years and for those aged 75 years or older, respectively. Risk differences were 187 events per 1000 (95% CI: 159-206) for patients with minor (NIHSS≤ 4) AIS and 32 events per 1000 (95% CI: 10-58) for those with moderate-to-severe AIS.

**Conclusions:** Initiating inpatient benzodiazepines within three days of AIS is associated with an elevated 10-day risk of falls or fall-related injuries, particularly for patients aged 65 to 74 years and for those with minor strokes. This underscores the need for caution with benzodiazepines, especially among individuals likely to be ambulatory during the acute and sub-acute post-stroke period.

## INTRODUCTION

Benzodiazepines are commonly employed in acute stroke care for purposes ranging from periprocedural sedation to short-term management of in-hospital complications such as insomnia, depression, and anxiety. With rising stroke incidence for patients over 65 years of age and the alarming rates of post-stroke depression in this age group, the potential for expanded use of benzodiazepines may give rise to unintended consequences ^1,2^.

Specifically, medical governing bodies have long advised against the long-term utilization of benzodiazepines in individuals older than 64 years due to increased vulnerability to adverse effects, such as somnolence and falls ^3^ . Moreover, risk is greater for older adults on polypharmacy regimens and those subject to acute brain injuries or enduring common post-stroke comorbidities such as gait disorders and depression. ^4,5^ Among AIS survivors, falls or fall-related injuries are primary drivers of emergent medical care utilization and loss of functional independence, manifesting at an annual incidence rate of up to 70% with a 4-fold higher fracture rate when compared to the general population ^6^ .

While risks associated with long-term benzodiazepine use are well established, avoidance is yet to be reflected in clinical practice and there remain conflicting views on whether short-term use carry similar risks^7–9^. Perhaps as a result, guidelines have failed to specifically address this issue, and some physicians dispute whether short-term prescriptions would represent a real public health problem ^3,10,11^.

Implementation of randomized clinical trials to determine the safety of short-term benzodiazepine usage during the acute AIS recovery phase is practically difficult and ethically challenging. Given the context of safety warnings associated with benzodiazepine use, the debatable safety of short-term administration, and the variability in initiation patterns, we employed a range of novel analytical methods to investigate whether early use of inpatient benzodiazepines within three days of an AIS, a threshold chosen to denote short-term inpatient use, results in net harm for adults 65 years and older ^8,10,12–16^. To mitigate the influence of confounding and selection biases in the context of non-randomized studies, we used a target trial methodology to emulate a pragmatic randomized clinical trial to assess the effect of short term inpatient benzodiazepine use on falls or fall related injuries (FRIs) for this population.

## METHODS

### Study Design and Treatment strategies

Specifically, the target trial would involve random assignment of eligible patients upon admission for AIS to one of two arms: i) treatment with benzodiazepines within the window extending from AIS admission to the third day post-admission or discharge date, should discharge happen before three days; ii) a control arm in which no benzodiazepines were given within the defined window extending from AIS admission to either the third day post-admission or discharge, whichever occurred first.

We assessed the primary outcome of the occurrence of a fall or FRI during a follow-up period spanning 10 days starting from the day of hospital admission. Further details regarding the observational analysis proposed to emulate this target trial can be found in **Table 2**.

### Data Sources

We leveraged the American Heart Association’s Get With The Guidelines (GWTG) Stroke Registry to identify eligible patients ^17,18^. The GWTG collected patient sociodemographic, health history, and clinical data detailing the stroke admission including stroke severity assessment defined by the validated Stroke Severity scale, NIHSS ^19,20^.

Once eligible patients were identified, we created a separate dataset by linking GWTG data to each patient’s electronic health record (EHR) from the Mass General Brigham (MGB) Healthcare System, thereby obtaining additional demographic, clinical, and health care utilization data, including diagnoses, procedures, outpatient, and inpatient drug administration ^21^. Data quality checks were done for each patient discharged from MGB with a stroke diagnosis before submission to the GWTG Registry ^17^.

This study was approved by the institutional review board of Massachusetts General Hospital, and informed consent was waived. The data that support the findings of this study are available from the corresponding author upon reasonable request and institutional approval.

### Study Population and Eligibility Criteria

We identified 3,532 patients aged 65 years and older who were admitted for AIS between January 1, 2014, and June 28, 2021, and had no recorded diagnosis of AIS in the prior 12 months. We excluded 35 patients without demographic information and 10 patients with missing encounter records. We excluded another 269 patients without a National Institutes of Health Stroke Scale (NIHSS) score recorded at admission. This enhanced the selection of AIS patients first seen at MGH for AIS symptoms because patients with missing NIHSS values on admission were typically transferred from another hospital one or more days after the initial cerebrovascular event^13^. We also excluded patients with one or more recorded prescriptions of benzodiazepines within the 3-month period before admission. The final eligible sample consisted of 3,059 patients (**Figure 1**).

**Figure 1.**
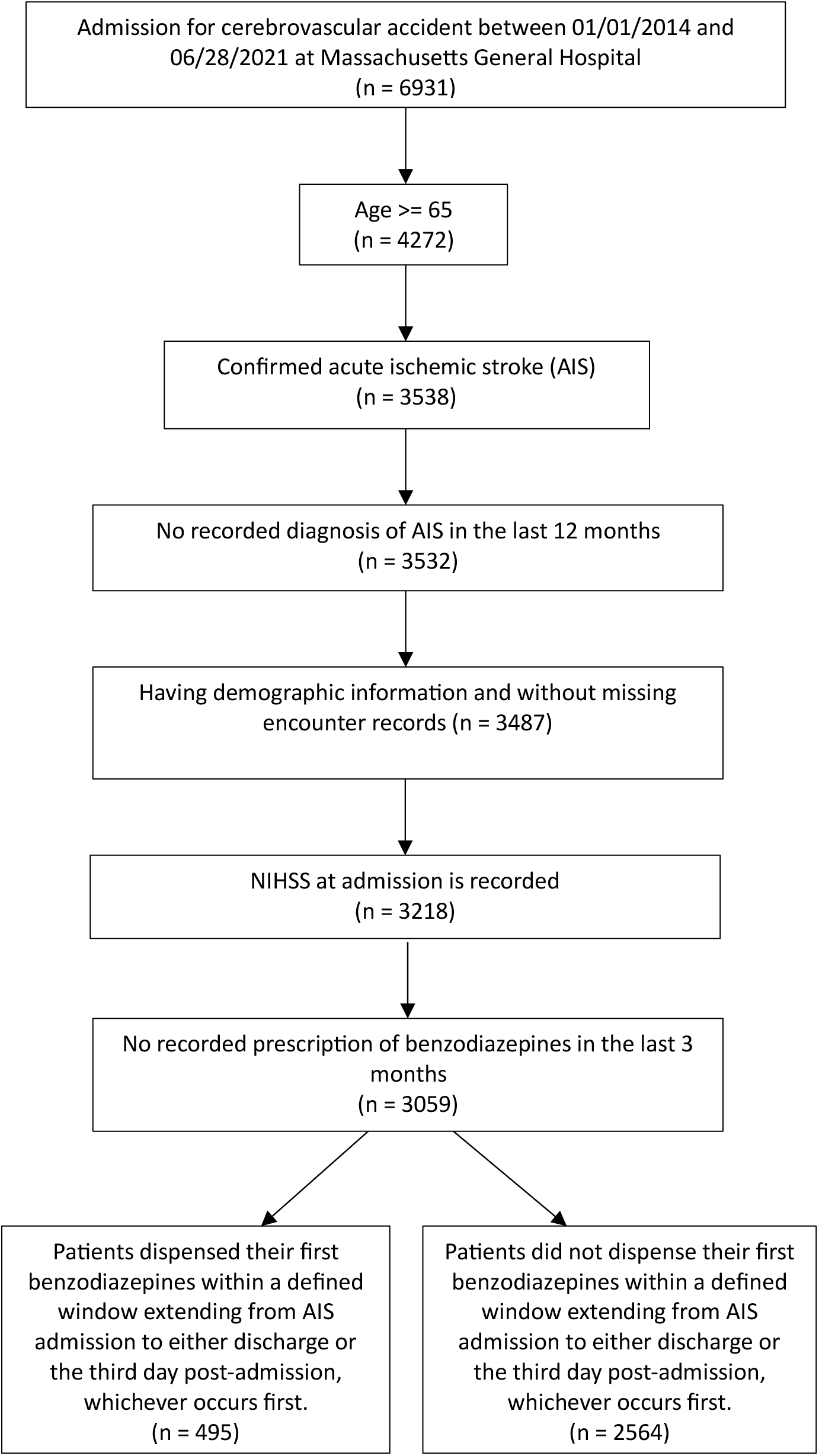
Selection of eligible patients with new acute ischemic stroke (AIS) ≥65 years, 2014-2021. Legend: Describes the sampling process that resulted in a sample of 3,059 patients, including patients ≥ 65 years at the time of new acute ischemic stroke admission, with available data in the electronic health record system and who had not received benzodiazepine in the three months before admission.

### Benzodiazepine Exposure and Treatment Assignment Procedures

In the target trial emulation, we obtained information on benzodiazepine use from inpatient and outpatient pharmacy data. The list of benzodiazepines used can be found in **Table S1** in the Supplementary Materials.

Within the context of the specified target trial, analysis of the available data requires consideration of immortal time bias and confounding bias ^22^. The former potentially arises since study entry (AIS admission) and benzodiazepine initiation could have occurred at different times, while the latter potentially arises since treatment assignment in the study sample was not actually randomized. To address these issues, we follow the approach of Hernàn and colleagues, which initially involves creating exact copies, or clones, of the patients ^15,16^. Subsequently, each clone is “assigned” to the treatment arm not received by the corresponding patient. This results in an effective doubling of the size of the dataset and ensures that potential baseline confounders are perfectly balanced between the two arms. To acknowledge the artificial doubling of the sample, person-time for each patient/clone is censored if and when the actual received treatment deviates from assignment. Consider, for example, a patient who received benzodiazepine during the exposure period. Their clone would have been initially “assigned” to the control arm, with their person-time censored at the time benzodiazepine by the patient was initiated ^15,16,23^. This approach ensures that eligibility, start of follow-up, and assignment to a treatment strategy are aligned and that patients can be represented in both arms until treatment can be determined ^24^. As such, baseline confounding bias has been addressed. We provide details of the cloning method in **Figure S1** and “Technical Details” in the online Supplementary Materials.

### Covariate Adjustment to Account for Artificial Censoring

Although the cloning method ensures that the baseline confounding factors are balanced, the artificial censoring itself may introduce time-dependent selection bias which can be addressed via inverse-probability of censoring weighting (IPCW) ^14,25,26^ . As a measure of stroke severity at baseline, we chose the NIHSS, a summary severity measure that is strongly associated with drug initiation ^19,20^. Importantly, NIHSS was reliably assessed, measured, and documented upon hospital admission (study time zero), making it an ideal baseline measure for use in the weights. We considered those with an NIHSS score ≤ 4 to have a minor AIS and those with an NIHSS score > 4 to have a moderate-to-severe AIS. We also used age and time-varying characteristics including time post-AIS and daily neurophysiology monitoring with an electroencephalogram ^13^. Technical Details in the Supplementary Materials provides the steps to perform the IPCW method.

### Follow-up and Outcome Measures

Patients were followed from AIS admission to the earliest of the following events: the date of fall or FRI or right censoring, which includes death (as recorded on EHR and/or GWTG registry), loss to follow-up (date of last use of EHR), or end of study (i.e., 10 days post admission including the admission day).

To accurately detect falls or fall-related injuries (FRIs) from various unstructured clinical notes— including admission, inpatient encounter notes and discharge summaries—we employed a validated Natural Language Processing (NLP) model, RoBERTa. This model was applied daily to all notes for each patient in our study sample, determining whether the patient experienced a fall or FRI on that particular day. This process was repeated for each day of the observation period. The model demonstrated high accuracy, with a precision of 0.90 (range 0.88-0.91), recall between 0.90 and 0.93, and an F1 score of 0.90 (range 0.89-0.92) ^27^. Additionally, it showed excellent Area Under the Receiver Operating Characteristic (AUROC) and Area Under the Precision-Recall (AUPR) curves, with scores of 0.96 (range 0.95-0.97) ^27^. Additional detail is available in supplementary text.

We extracted death dates from the EHR Demographics data file (Death Master File). Given that MGB updates deaths of patients monthly with data from the Social Security Administration, deaths were captured even if the patient was transferred into a nursing home or another non-MGB facility.

### Statistical analysis

We describe the characteristics of eligible patients stratified by benzodiazepine initiation strategy in **Table 1**.

**Table 1.**
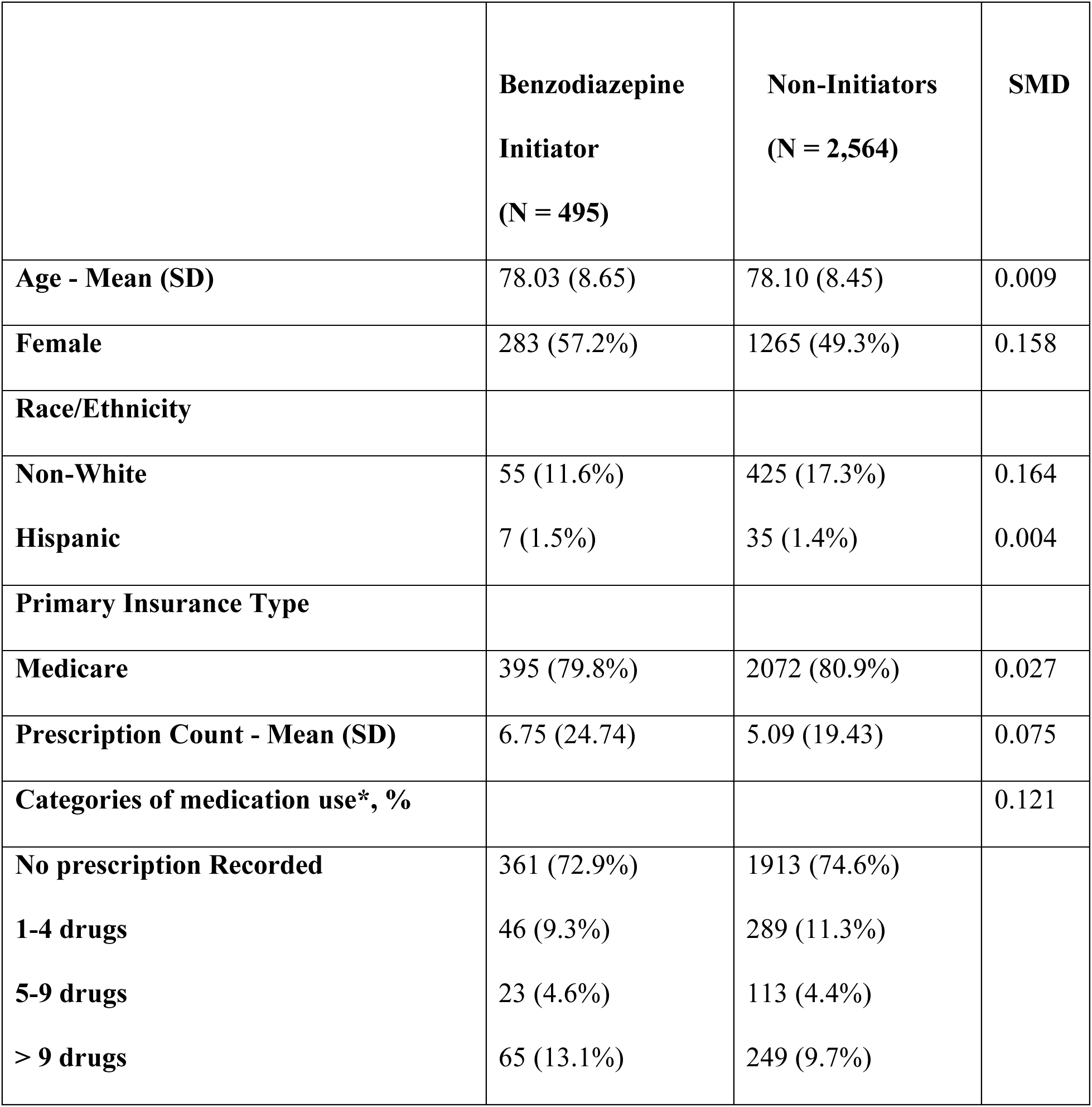
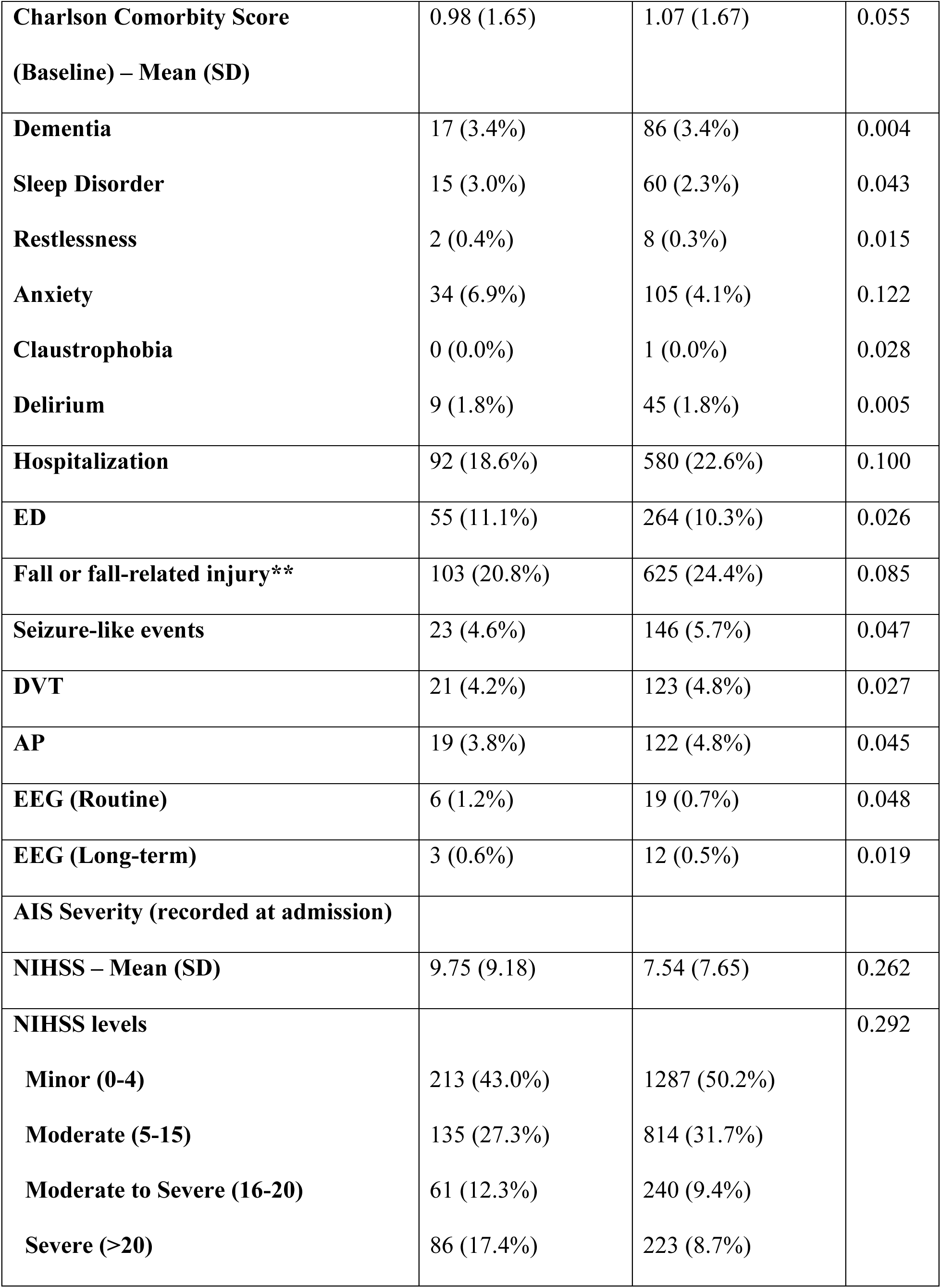
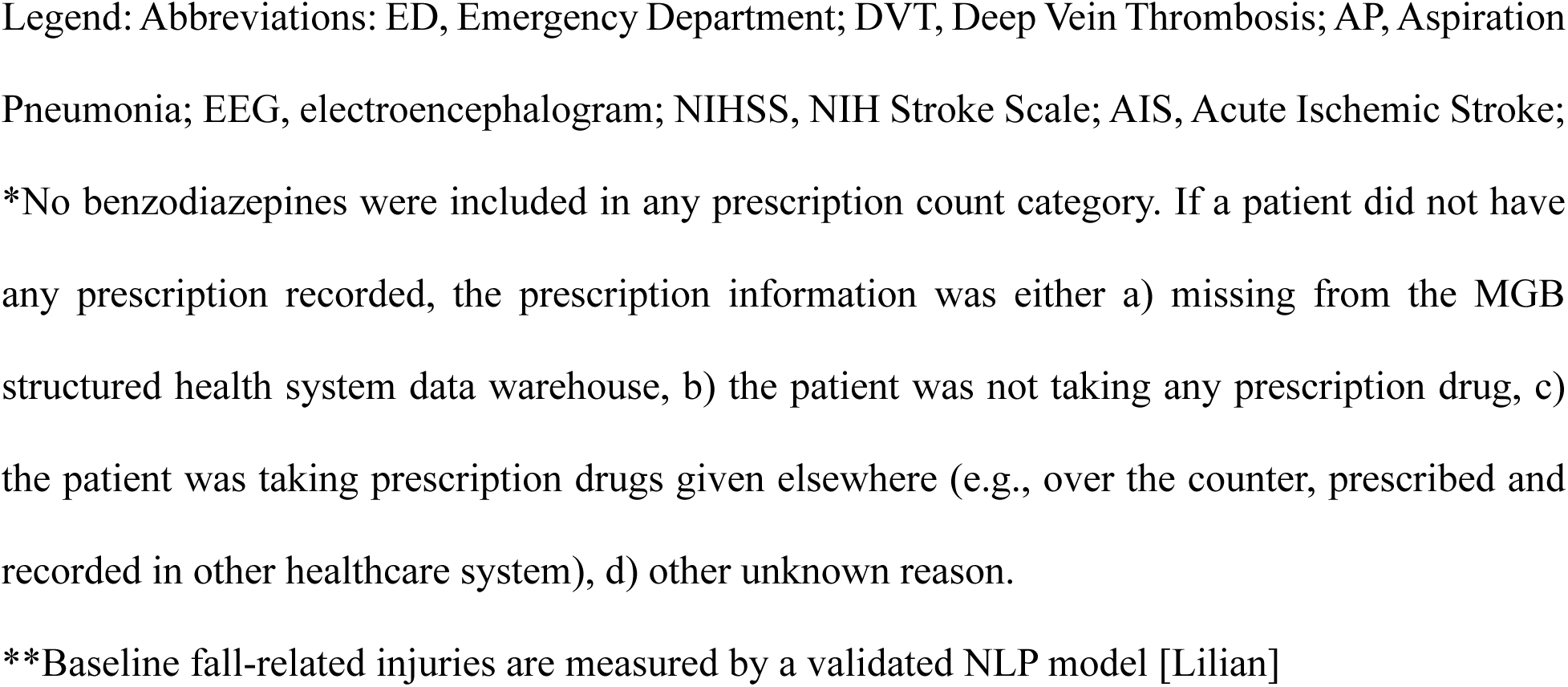
Characteristics of patients stratified by benzodiazepine initiation within a defined window extending from AIS admission to either discharge or the third day post-admission versus non-benzodiazepine initiation within a defined window extending from AIS admission to either discharge or the third day post-admission.

**Table 2.** Description of a target trial and the corresponding observational study.

| Target trial | Emulated trial (observational study) |
| --- | --- |
| Eligibility criteria |  |
| Admission for cerebrovascular accident between 01/01/2014 and 06/28/2021 at Massachusetts General Hospital | Same |
| Age $\geq 65$ | Same |
| Confirmed Acute Ischemic Stroke (AIS) | Same, and exclude those without NIHSS recorded at hospital admission |
| No previous history of AIS in the last 12 months | No recorded diagnosis of AIS in the last 12 months |
| No use of benzodiazepines in the last 3 months | No recorded prescription of benzodiazepines in the last 3 months |

| Treatment strategies |  |
| --- | --- |
| Treatment arm: Initiate benzodiazepine**** within a defined window extending from AIS admission to either discharge or the third day post-admission, whichever occurs first.<br>Control arm: Refrain from giving benzodiazepines within the defined window extending from AIS admission to either discharge or the third day post-admission, whichever occurs first | Same |
| Assignment procedures |  |
| Open label, randomized treatment assignment | Emulated randomization* |
| Outcomes |  |
| Time to fall or fall related injury from the day of AIS admission | Same. Time to fall or fall related injury as determined by NLP algorithms |
| Follow-up |  |
| Starts at randomization (at admission)<br>Ends at date of fall or fall-related injury, death, loss to follow-up, or end of study | Starts at admission.<br>Ends at date of fall or fall-related injury, death (as recorded in EHR and/or GWTG registry), loss to follow-up (date of last use of EHR**), or end of |
| (i.e., 10 days post admission including the admission day); whichever occurs first | study (i.e., 10 days post admission including the admission day); whichever occurs first |
| Causal contrast |  |
| Intention-to-treat effect | Observational analog of intention-to-treat effect*** |
| Statistical analysis |  |
| Intention-to-treat effect analysis of time to fall or fall-related injury, accounting for censoring. | Same, additionally accounting for baseline confounding. |

Legend: Abbreviations: NIHSS, NIH Stroke Scale; AIS, Acute Ischemic Stroke; GWTG, Get-with-The-Guidelines Stroke Registry; EHR, electronic health record; IPCW, Inverse Probability of Censoring Weights.
\*Emulated randomization – Emulated randomization by balancing confounders using IPCW for treatment selection. Please see additional technical details on methods used to address immortal time bias.
\*\*Date of last use of EHR – see technical details on follow-up data quality and quantity.
\*\*\*Analysis can be consistent with “intention-to-treat” (with respect to treatment assignment as a strategy [treat vs not]). However, the effect can also be analogous to the “per-protocol” (when treatment strategy is considered a “protocol” that must be followed overtime [the grace period]).
\*\*\*\*Benzodiazepine: listed in **Table S1** of the Supplementary Materials.

To evaluate the effect of benzodiazepine initiation within the defined exposure period of post-AIS on 10-day falls or FRIs, we estimated the survival probability of a fall or FRI using model-based predictions. To do so, as indicated above, we expanded the original data by creating clones, censored the clones as previously described, and weighted the expanded data using stabilized IPCWs to account for artificial censoring. The models for IPCW were pooled linear logistic regressions over person days including age, NIHSS, daily use of neurophysiologic monitoring (EEG), days post-AIS admission, and potential interactions between them, separately for the two treatment strategies. For comparison, we estimated the unadjusted Kaplan-Meier and a cubic spline model-based survival probabilities of falls or FRIs in the 10-day period using the cloned data set. See Figure S1 in the Supplementary Materials.

Using the expanded weighted data, we fitted a pooled logistic regression model for falls or FRIs as a function of the following covariates: treatment strategy, time (measured in days post-AIS admission), a quadratic term of time, and an interaction term of treatment strategy and time to allow for time-varying effects. We obtained estimated survival probabilities for each day under each treatment strategy and estimated the 10-day fall or FRIs risk difference. Additional analysis considering the potential different inpatient and outpatient risks of falls or FRIs is provided in the “Additional Analysis” section in the Supplementary Materials.

Additionally, whenever the date of benzodiazepine prescription coincided with the day of discharge we deferred to the inpatient/outpatient marker in the dataset to determine exposure status. Such instances occurred for 24 individuals.

We used the bootstrap method based on 500 re-samplings to obtain 95% confidence intervals, accounting for inflation of the sample size by cloning. See Technical Details in the Supplementary Materials for model specifications and additional statistical analysis details.

### Missing Data and Pre-planned Stratified Analysis

We examined patterns of missing data for all relevant variables to confirm that the analysis had minimal missing information with less than 5% missingness on all variables ^10,13^. The sedating effects of benzodiazepines on ambulation dexterity might be more pronounced among AIS survivors who can walk during the early days of recovery (e.g., those with mild AIS) and are therefore at risk for falls or FRIs. Additionally, benzodiazepines may be more detrimental to older patients. Therefore, we repeated the analyses above, stratifying by age and NIHSS categories. We also conducted two secondary analyses related to the length of follow-up, specifically at 8 and 30 days post admission; see the “Additional Analysis” section in the Supplementary Materials.

## RESULTS

### Study population characteristics

Among patients aged 65 years and above, 3,059 were eligible for the emulated target trial. Of those, 495 (16%) initiated a benzodiazepine within the third day post-admission or discharge, whichever occurred first. **Table 1** describes demographic and clinical characteristics of patients stratified by benzodiazepine initiation strategies. The most frequently prescribed benzodiazepine was lorazepam (86%, **Table S2**)

### Outcome: Falls or fall-related injuries

We obtained standardized survival functions averaging over the distribution of covariates for the selected population after cloning by benzodiazepine initiation strategies for all 3,059 eligible patients during the first 10 days post-AIS admission (**Figure 2**). After target trial emulation with cloning, the unadjusted Kaplan-Meier 10-day risk of falls or FRIs was 530 events per 1000 patients among those under the benzodiazepine initiation strategy and was 561 events per 1000 patients among those under the non-initiation strategy.

**Figure 2.**
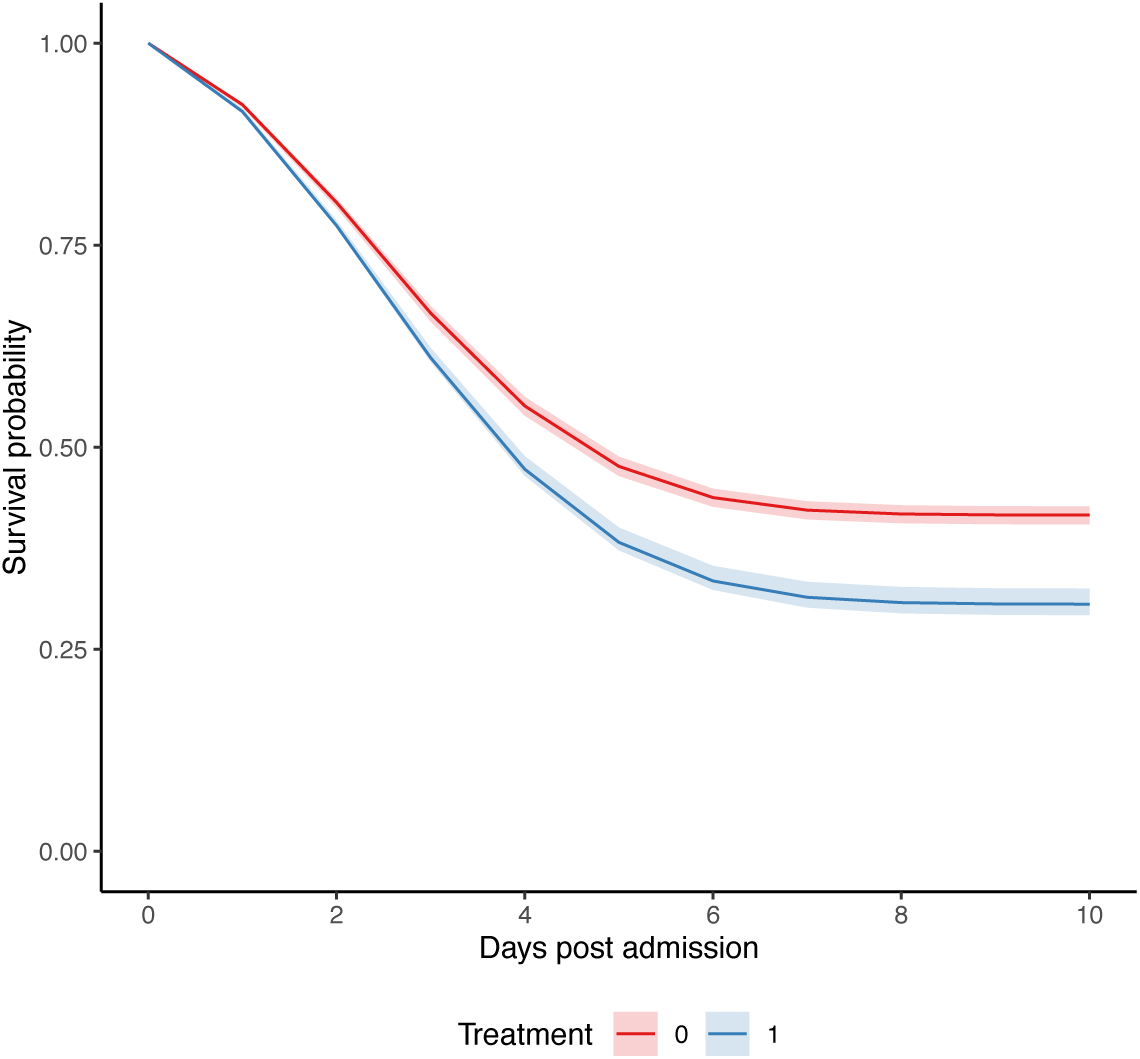
Standardized survival curves by benzodiazepine initiation strategy. pooled logistic regression survival curves for all patients. Blue: Strategy for benzodiazepine initiation within three days post-AIS admission (or discharge). Red: Strategy for no initiation of benzodiazepine within three days post-AIS admission. Shaded areas: 95% confidence intervals constructed using bootstrap with 500 replications.

After target trial emulation with cloning and adjustment for artificial censoring, standardized 10-day risk of falls and FRIs was 694 events per 1000 (95% CI: 676-709) for those under the benzodiazepine initiation strategy and 584 events per 1000 (95% CI: 575-595) for those under the non-initiation strategy, resulting in a risk difference of 110 events per 1000 patients (95% CI: 89-125).

Among AIS patients aged 65 to 74 years and those aged 75 years or older, the 10-day risk differences were 142 events per 1000 patients (95% CI:111-165) and 85 events per 1000 patients (95% CI: 64-107), respectively (**Figure 3**). Among AIS patients with minor AIS (NIHSS ≤ 4) and moderate-to-severe AIS (NIHSS > 4), risk differences were 187 events per 1000 patients (95% CI: 159-206) and 32 events per 1000 patients (95% CI: 10-58), respectively (**Figure 4**). The standardized 10-day risks of falls or FRIs stratified by age and NIHSS are displayed in **Table S3** in the Supplementary Materials.

**Figure 3:**
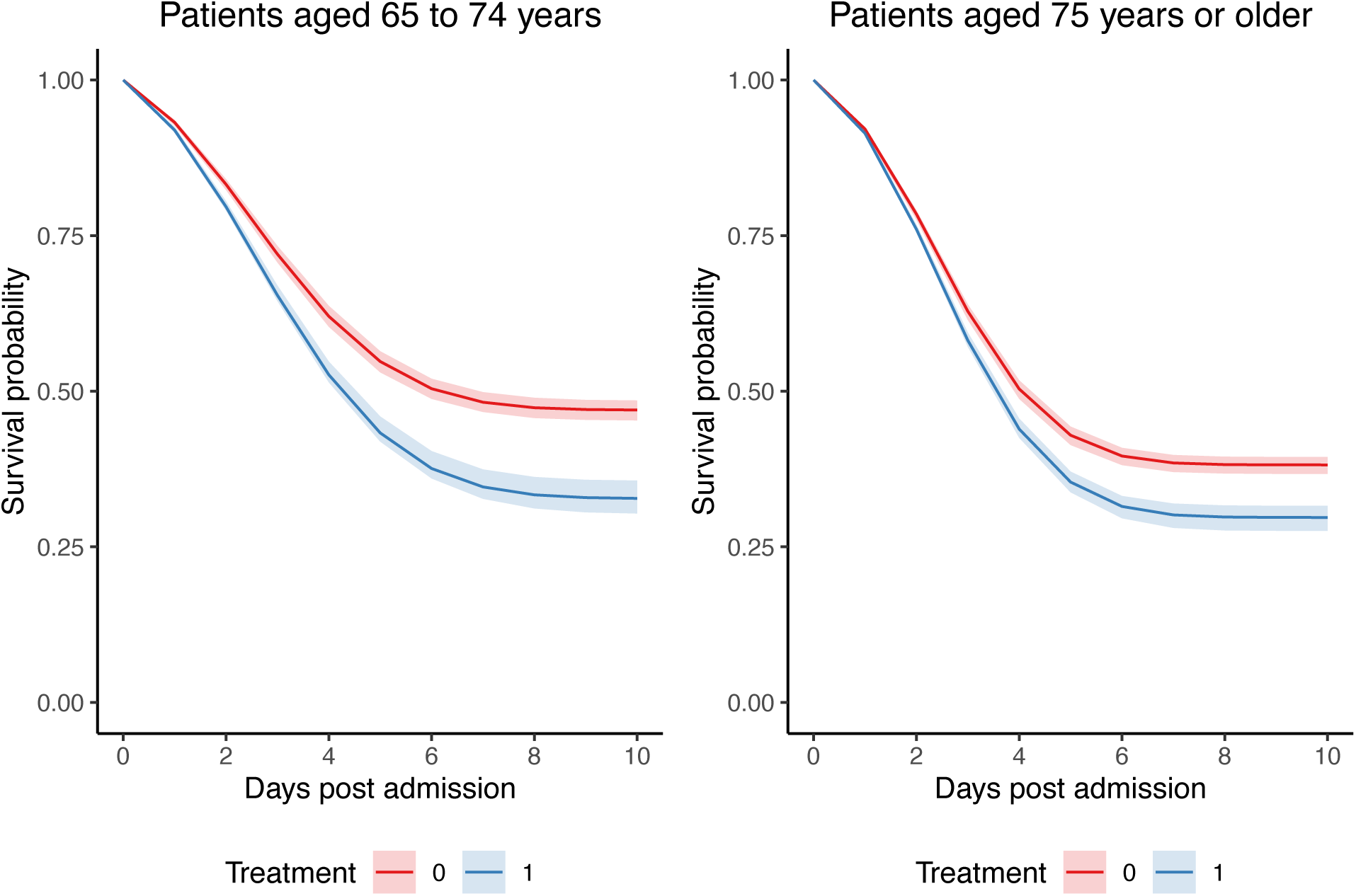
Standardized Survival Curves by Benzodiazepine Initiation Strategy Across Categories of Age. Blue: Strategy for benzodiazepine initiation within three days post-AIS admission (or discharge). Red: Strategy for no initiation of benzodiazepine within three days post-AIS admission. Shaded areas: 95% confidence intervals constructed using bootstrap with 500 replications.

**Figure 4:**
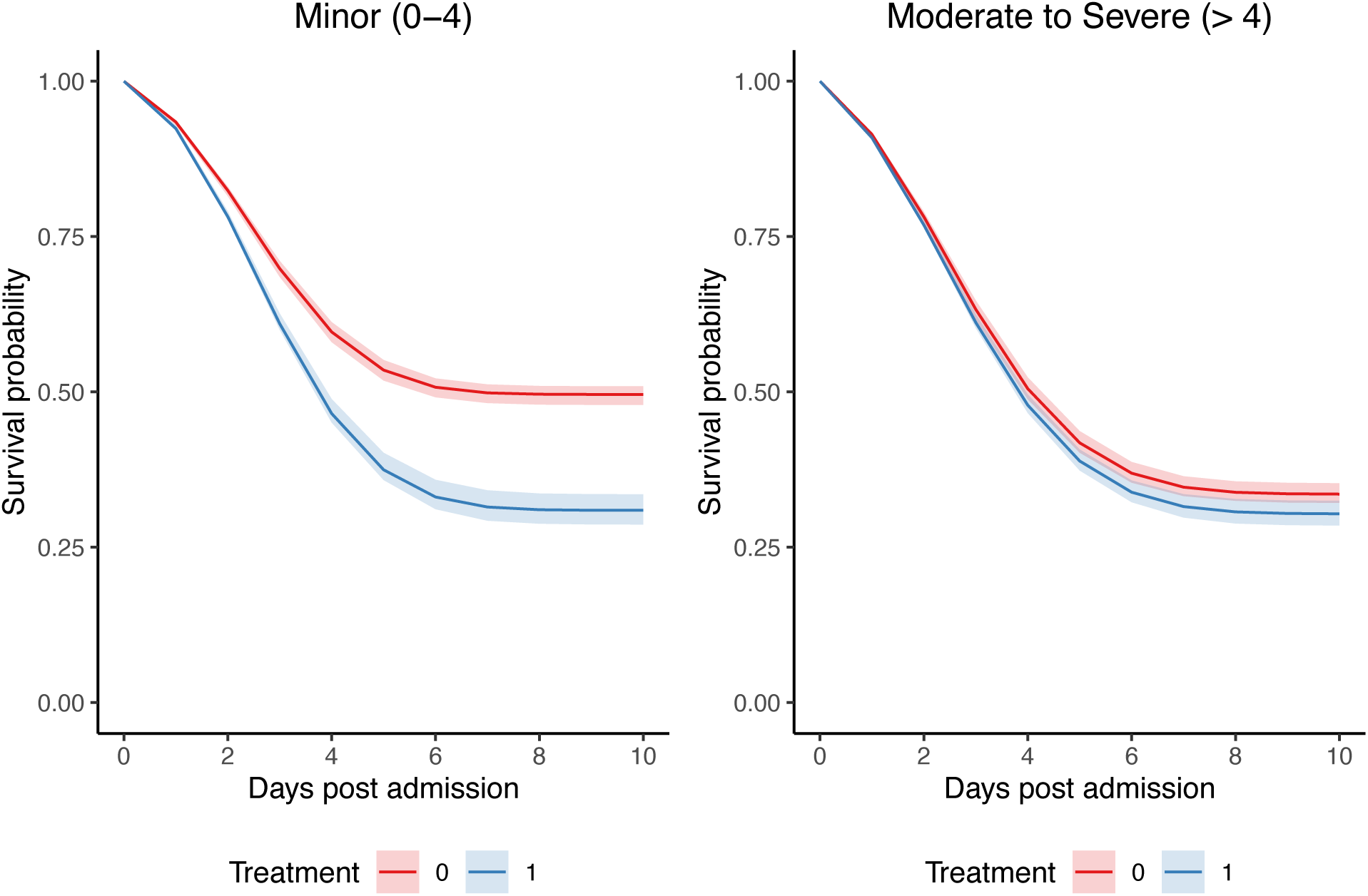
Predicted pooled logistic regression survival curves stratified by NIHSS scores. Blue: Strategy for benzodiazepine initiation within three days post-AIS admission (or discharge). Red: Strategy for no initiation of benzodiazepine within three days post-AIS admission. Shaded areas: 95% confidence intervals constructed using bootstrap with 500 replications.

Repeating the analysis with 8-day and 30-day follow-up windows yielded similar results, consistent with an excess of standardized risk of falls or FRIs associated with initiation of benzodiazepines. Considering that the inpatient and outpatient risk of falls or FRIs could be different, we repeated the analysis by adding a time-varying discharge indicator in the outcome model with 10-day follow-up window. The results appeared to be consistent with the main result in the manuscript. See Section “Additional Analysis” in the Supplementary Materials. The Supplementary Text provides the Statistical Code used to conduct the main analysis.

## DISCUSSION

Our study emulated a hypothetical randomized trial of inpatient benzodiazepine use in older adults during the acute post-stroke recovery period to assess incidence of falls or FRIs. While some argue judicious, short-term use of benzodiazepines carries neglectable, limited risks, we estimated that there were 110 more events per 1000 patients in the benzodiazepine initiator strategy group^28^. The tight confidence intervals and the consistency of results using multiple alternative follow-up periods points to a causal effect.

Furthermore, we observed higher estimates of falls or FRIs risks among patients aged 65 to 74 years; they had an excess rate of 142 events per 100 patients. Higher risks were also seen among those with minor AIS as defined by an NIHSS scores less ≤ 4. This contrasts with recent population level analysis that could only perceive increased fall risks in patients greater than 80 years with Medicare’s recent benzodiazepine coverage expansion ^29^. While we observed greater risk among mild stroke cases, this can largely be attributed to differences in ambulatory status. Therefore, our results align with the expectation that benzodiazepines confer increased fall risk on older patients with reduced dexterity and mobility that have sustained any forms of brain injury yet remain able to ambulate (and fall). ^4^

This is particularly concerning as guidelines have long been ambiguous in condemning short-term use for purposes such as periprocedural sedation and severe generalized anxiety in inpatient care ^3,30^. While no studies have specifically addressed fall risk increase with short lived exposures, it was noted that even fast acting benzodiazepines can lead to higher number of falls in the general elderly population ^31,32^. Additionally, tolerance is largely restricted to repeated exposures, which lends biological plausibility to the concern that new users may be at even higher risk ^33^.

A major strength of this study revolved around our ability to capture falls or FRIs from unstructured data disposing of the need to rely on ICD codes alone. Such events have historically lacked comprehensive documentation in structured datasets, thereby limiting strategies to research validity ^34–37^. To address this concern, we employed a validated Natural Language Processing (NLP) model designed for discerning falls or FRIs within unstructured clinical notes ^27,38^. Additionally, we had minimal to zero missing data on pertinent variables which is both due to refined patient selection criteria and detailed data registry.

### Limitations

#### Residual Confounders

The unadjusted analysis yielded results contrary to those observed in the standardized analysis, suggesting the presence of multiple confounding factors. Intubation for instance would be an indication of benzodiazepines for more severe patients that essentially prevents falls for patients who are sedated and don’t have sufficient mobility to ambulate and fall. With the use of NIHSS admission score we determined stroke severity and were able to perform stratified analysis based on overall clinical picture and its mobility implications. In a similar fashion, specific frailty markers including walking speed and gait metrics (e.g. time to stand) or overall strength and energy would have provided further insights for this population but were unavailable.

As many of the factors associated with receipt of benzodiazepine and falls were highly correlated with baseline stroke severity and comorbidities, it is possible that adjustment for NIHSS scores and the elicited comorbid conditions is balancing for other unmeasured confounders. Our assumption therefore is that after accounting for such, residual confounding by unrecorded measures is minimal.

#### Imperfect Exposure and Outcome Measures

Although refined, the output of the NLP-based strategy to measure the outcome of interest (falls or fall-related injuries) included the date in which the documentation occurred but not necessarily the specific dates in which the actual events occurred. For instance, it is possible that if a fall occurred in one night it could have been documented only in the next day or at the discharge notes. This may introduce misclassification bias with respect to time to event, but it is expected to have occurred randomly (independent of treatment strategy). The issue of documentation of events that occurred remotely is still possible but less likely in this study, as we selected a sample with an acute ischemic event and focused analysis to the inpatient context where the documentation of remote acute events is less likely to occur. An ideal measure of outcome would have integrated both time in which events were documented and time in which they actually occurred, which could be the focus of future NLP validation studies.

#### Generalizability

In this single-center study, our data was collected from a large academic institution with a predominantly white, non-Hispanic, and insured patient population which yielded patients with larger recorded health care system utilization. This favored our internal validity and allowed for better confounder control at the expense of generalizability and representativeness.

## CONCLUSIONS

This study examined the likelihood of 10-day falls or FRIs risk associated with the initiation of inpatient benzodiazepine in patients ≥ 65 years within three days after an AIS. After standardization, we found a greater likelihood of a fall or FRI within 10 days of admission associated with inpatient benzodiazepine being initiated within three days of an AIS. This reflects the idea that benzodiazepine associated fall risks are also significant in the short-term and highlight the need for new guidelines.

## Data Access and Responsibility

Shuo Sun had full access to all the data in the study and takes responsibility for the integrity of the data and the accuracy of the data analysis. All authors certify that they had sufficient access to the data to verify the scientific integrity of the manuscript.

